# A nine-year investigation of industry payments to emergency physicians in the United States between 2013 and 2021

**DOI:** 10.1101/2023.07.24.23293098

**Authors:** Anju Murayama

## Abstract

**Objectives:** To examine the characteristics and trends in the industry payments to emergency physicians since the inception of the Open Payments Database in 2013 and the COVID-19 pandemic in 2020.

**Methods:** Using the Open Payments Database between August 2013 and December 2021, this population-based cohort study examined all research and general payments made by the healthcare industry to emergency physicians registered in the National Plan and Provider Enumeration System in the United States. We performed descriptive analyses on payment data and generalized estimating equations for payment trends.

**Results:** Among 50,483 active emergency physicians, 28,678 (56.8%) accepted a total of $457,640,796.73 payments from the healthcare industry between 2013 and 2021. 56.6% and 1.3% of all emergency physicians received general and research payments, respectively. 20.8% ($94.98 millions) of overall industry payments were general payments. Median general and research payments per-physician (interquartile range) were $133.21 ($44.78–$355.77) and $62,842.97 ($10,320.00–$273,285.28), respectively. Top 1% of emergency physicians received 86.2% of overall general payments, respectively. The number of physicians receiving general payments decreased by 2.9% (95% CI: −3.2 to −2.5, p<0.001) annually between 2014 and 2019 and 47.8% (95% CI: −49.8 to −45.6, p<0.001) in 2020. Although there were no significant changes in research payments before COVID-19 pandemic, the research payments significantly increased by 69.4% (95% CI: 28.9–122.7, p<0.001) in 2021 than those in 2020.

**Conclusions:** Majority of emergency physicians accepted general payments from the healthcare industry, but the number of emergency physician accepting the general payments significantly decreased since the inception of Open Payments Database.

## Introduction

Financial relationships between physicians and the healthcare industry are widespread for research and non-research purposes in the United States (US).[1,2] In response to increased public demand for greater transparency and revelation of their negative impact on physicians’ clinical practice and research, the Physician Payments Sunshine Act was enacted in 2010. It legally mandates all pharmaceutical and medical devices manufacturers to disclose their payments to physicians on a federal transparency database, the Open Payments Database since 2013.[1,3,4]

Open Payments Database contains comprehensive physician-industry financial relationships in the US, including emergency physicians. Two previous studies using the Open Payments reported physician-industry financial relationships in the field of emergency medicine. Fleischman et al. first reported about one third of emergency physicians received general payments in 2014.[5] Later, Niforatos et al. documented that more than one third of emergency physician still received general payments from the healthcare industry in 2017.[6]

Although previous studies on emergency physicians in the US exists, little is known about the longitudinal trends in the industry payments to emergency physicians since the inception of the Open Payments Database in 2013. Additionally, the sudden onset of COVID-19 pandemic significantly impacted physician activities and industry marketing strategies in the US, [7,8] while there was no document concerning the emergency physician-industry relationships during the COVID-19 pandemic. This study conducted the first longitudinal analysis with large sample size spotlighting the whole picture of the magnitude, prevalence, and trends in the physician-industry financial relationships since the inception of the Open Payments Database and the COVID-19 pandemic in the US in the field of emergency medicine.

## Methods

### Study design and setting and Selection of Participants

This cross-sectional analysis evaluated the magnitude, prevalence, and trend in both general and research payments to emergency physicians since the inception of the Open Payments database in 2013. This study defined emergency physician as a physician whose primary specialty was categorized as emergency medicine and entailed subspecialties in the National Plan and Provider Enumeration System (NPPES) database.

All emergency physicians who registered to the NPPES before August 1, 2013, were included in this study. As this is longitudinal analysis aiming to examine the trends in industry payments, emergency physicians who newly activated or deactivated after August 1, 2013, were excluded from the study sample.

### The Open Payments Database and data collection

The payments and relevant data were collected in the following steps. First, all profile data of emergency physicians were extracted from the NPPES database. Second, the extracted NPPES dataset were matched with the Open Payments physician profile database using the National Provider Identifier number. Third, all general payments made to the emergency physicians between August 2013 and December 2021 were extracted from the Open Payments payment database using the covered recipient numbers. Finally, for the research payments, all research payments of which the principal investigator’s NPPES primary specialty was emergency medicine were collected. This method includes indirectly paid research payments via third parties (e.g. teaching hospitals and universities), preventing underestimation of the magnitude and prevalence of research payments, as shown by previous studies.[9,10]

### Outcome and statistical analyses

This study conducted descriptive analysis on the extracted payments data on individual physician level. Per-physician payments were calculated only among physicians receiving payments each year and overall, as performed in other previous studies.[1,3,11-13] Per-physician payments were calculated based on total amounts of payments divided by the number of principal investigators in a research payment. To examine the concentration of general and research payments per physician, the share of payments by specific proportions of physicians and the Gini index were calculated. The Gini index ranges from 0 indicating complete equality to 1 indicating complete inequality.[12,13]

Additionally, the trends in general and research payments were evaluated by the population-averaged generalized estimating equations (GEE) model with panel-data of annual payments per physician clustering each physician level, as noted previously.[7,8,13,14] To take in account in the influence of the COVID-19 pandemic since 2020, interrupted time series (ITS) analysis were employed. The ITS analysis with robust negative binomial regression GEE model for trend in payments per physician and ITS analysis with modified robust log-linked regression GEE model with Poisson distribution for trend in the number of physicians receiving payments between 2014 and 2021 were applied, as the payments were highly skewed.[13,14] Payment data in 2013 was excluded from the trend analysis, as the payment data in 2013 was partially disclosed between August and December 2013. The research payment data were rounded down as a unit of $10,000 to stabilize the ITS GEE model. The study also conducted ITS analysis excluding the payments for acquisitions, debt forgiveness, long-term medical supply or device loans, ownership interests, and royalties and licenses, because there were substantial amounts of general payments for royalties and license in several years or these payment categories were newly-added in 2021. All data extraction and data analyses were performed with Python 3.9.12 (Python Software Foundation, Beaverton, OR, US), Microsoft Excel, version 17.0 (Microsoft Corp., Redmond, WA, US), and Stata version 15 (StataCorp, College Station, TX, US).

### Ethical clearance

No institutional board review and approval were required for this study as all data used met the definition of freely available public data.

## Results

### Overview of industry payments to emergency physicians

There were 50,483 active emergency physicians as of August 1, 2013, in the NPPES database. Among 50,483 active emergency physicians, 28,678 or 56.8% of all emergency physicians received a total of 420,967 summing $457,640,796 from 1210 different companies between 2013 and 2021.

### General payments

As of general payments, 28,590 emergency physicians (56.6%) received 321,428 general payments totaling $94,977,251 (20.8% of overall industry payments) from 1145 different companies over the nine years. 16 (0.005%) general payments were under dispute and 15 (93.8%) were unchanged. Median payment was $17.32 (interquartile [IQR]: $13–$73), while average payment was $299 (standard deviation [SD]: $22,103). Median total payments per physician were $133 (IQR: $45–$356) and average payments were $3,322 (SD: $91,063). (Table 1) Median annual total general payments ranged from $31 (IQR: $16–$103) in 2020 to $54 (IQR: $19–$131) in 2014. Gini index for the total general payments per physician was 0.975, suggesting the only the very small number of emergency physicians received substantial amounts of general payments. Only top 1%, 5%, 10%, and 25% of all emergency physicians received 86.2%, 94.4%, 96.5%, and 98.9%, respectively. Gini index was 0.975 for per-physician general payments, showing that only the very small number of emergency physicians received substantial amounts of general payments.

**Table 1.** General payments to emergency physicians between 2013 and 2021.

| Variables | Program year |  |  |  |  |  |  |  |  | Overall |
| --- | --- | --- | --- | --- | --- | --- | --- | --- | --- | --- |
|  | 2013<sup>†</sup> | 2014 | 2015 | 2016 | 2017 | 2018 | 2019 | 2020 | 2021 |  |
| General payments |  |  |  |  |  |  |  |  |  |  |
| Total payments |  |  |  |  |  |  |  |  |  |  |
| Payment amounts, \$ | 3,802,347 | 10,299,971 | 11,355,294 | 23,551,636 | 12,476,914 | 8,336,995 | 9,010,822 | 6,451,731 | 9,691,541 | 94,977,251 |
| Number of payment contracts, n | 17,023 | 41,903 | 48,400 | 48,813 | 43,546 | 36,957 | 35,858 | 20,325 | 28,603 | 321,428 |
| Number of physicians with payment, n (%) | 6,627 (13.1) | 10,433 (20.7) | 13,126 (26.0) | 12,418 (24.6) | 11,628 (23.0) | 10,343 (20.5) | 9,935 (19.7) | 6,486 (12.9) | 7,672 (15.2) | 28,590 (56.6) |
| Payments per physician, \$ | | | | | | | | | | |
| Median (IQR) | 36 (15–111) | 54 (19–131) | 52 (18–125) | 50 (19–125) | 50 (19–125) | 47 (18–129) | 54 (19–135) | 31 (16–103) | 43 (18–125) | 133 (45–356) |
| Average (SD) | 574 (5,910) | 987 (17,059) | 865 (9,879) | 1,897 (109,478) | 1,073 (21,653) | 806 (6,928) | 907 (7,396) | 995 (7,701) | 1,263 (13,214) | 3,322 (91,063) |
| Range | 1–259,567 | 0.01–1,431,778 | 1–591,106 | 3–12,149,520 | 1–2,116,142 | 2–299,999 | 1–281,792 | 2–213,163 | 1–579,350 | 1–14,265,662 |
| Gini index | 0.990 | 0.989 | 0.984 | 0.993 | 0.988 | 0.986 | 0.987 | 0.993 | 0.993 | 0.975 |

Abbreviations: interquartile range (IQR), SD (standard deviation)
<sup>†</sup> The payment data in 2013 were payments made to physicians between August 1 and December 30.

Table 2 describes the general payments by payment categories. Of 15 payment categories, payments for speaking fees other than continuous medical education (CME) purposes occupied 38.5% ($36,522,818) in monetary values, while only 841 (1.7%) emergency physicians received 9,101 (2.8%) payments. Meanwhile, only 0.2% (109) emergency physicians received 703 (0.2%) payments for CME speaking purposes totaling $1,521,937 (1.6%). Payments for food and beverage was the largest number of emergency physicians receiving payments and number of payments, with 27,966 (55.4% of all emergency physicians) and 274,678 (85.5% of overall). The payments with the second and third largest number of physicians receiving payments were education and travel payments with 2,964 (5.9%) and 1,543 (3.1%), respectively.

**Table 2.** General payments by categories.

| Payment categories | Payment amounts,<br>\$ (%) | Number of<br>payments, n<br>(%) | Number of<br>physicians accepting<br>payments, n (%) |
| --- | --- | --- | --- |
| Non-CME speaking<sup>a</sup> | 36,522,818 (38.5) | 9,101 (2.8) | 841 (1.7) |
| Consulting fees | 33,292,171 (35.1) | 11,901 (3.7) | 1,348 (2.7) |
| Food and beverage | 9,264,816 (9.8) | 274,678 (85.5) | 27,966 (55.4) |
| Travel and lodging | 6,314,702 (6.6) | 16,945 (5.3) | 1,543 (3.1) |
| Honoraria | 3,102,053 (3.3) | 1,672 (0.5) | 613 (1.2) |
| CME speaking fees<sup>b</sup> | 1,521,937 (1.6) | 703 (0.2) | 109 (0.2) |
| Royalty or license | 1,387,444 (1.5) | 92 (0.03) | 13 (0.03) |
| Current or<br>prospective<br>ownership or<br>investment interest | 1,249,771 (1.3) | 45 (0.01) | 18 (0.04) |
| Education | 617,592 (0.7) | 5,379 (1.7) | 2,964 (5.9) |
| Grant | 525,701 (0.6) | 43 (0.01) | 33 (0.1) |
| Long term medical<br>supply or device loan | 436,062 (0.5) | 21 (0.01) | 13 (0.03) |
| Acquisitions | 383,421 (0.4) | 4 (0.001) | 3 (0.03) |
| Gift | 341,173 (0.4) | 679 (0.2) | 415 (0.8) |
| Entertainment | 8,889 (0.01) | 159 (0.05) | 104 (0.2) |
| Charitable<br>contribution | 8,700 (0.01) | 6 (0.002) | 5 (0.01) |

<sup>a</sup>Non-CME speaking payments included “compensation for services other than consulting, including serving as faculty or as a speaker at a venue other than a continuing education program.”
<sup>b</sup>CME speaking payments included “compensation for serving as faculty or as a speaker for an accredited or certified continuing education program” (applicable to program years 2013-2020), “compensation for serving as faculty or as a speaker for an unaccredited and non-certified continuing education program” (applicable to program years 2013-2020), and “compensation for serving as faculty or as a speaker for medical education program” (applicable beginning with program year 2021 and subsequent program years).

### Research payments

Only 663 equivalent to 1.3% of all emergency physicians received 99,539 research payments summed $362,663,547 (79.2% of overall industry payments) from 272 companies between 2013 and 2021. No research payment was disputed. Per-physician research payments were $62,843 (IQR: $10,320–$273,285) in median and $547,004 (SD: $1,884,010) in average. Median annual total general payments ranged from $19,541 (IQR: $3,676–$95,671) in 2014 to $44,943 (IQR: $11,453–$200,961) in 2021.Gini index was 0.998 for per-physician nine-year combined research payments. Among 663 emergency physicians receiving research payments, 575 (86.7%) received $1,121 (IQR: $160–$7,491) in median total general payments, while emergency physicians without research payments received $131 (IQR: $44–$342) in median.

Of 99,539 research payments, only 786 research payments totaling $4.62 million, equal to 0.8% in the number of payments and 1.3% in monetary values of overall research payments, were made for preclinical research. Additionally, there were 5,579 research payments totaling $56,925,292 (15.7% of all research payments) related to 296 different registered clinical trials. Emergency physician principal investigators of a randomized, placebo-controlled trial assessing efficacy of bamlanivimab and etesevimab in mild to moderate COVID-19 patients (ClinicalTrials.gov Identifier: NCT04427501)[15] received the largest total research payments of $5,449,62, followed by a clinical trial assessing safety and efficacy of SARS-CoV-2 monoclonal antibodies (ClinicalTrials.gov Identifier: NCT04425629)[16] with $4,638,617 and a clinical trial assessing the efficacy of alone or in combination of SARS-CoV-2 monoclonal antibodies (ClinicalTrials.gov Identifier: NCT04634409)[17] with $4,021,590.

### Summary and Trends in annual general and research payments

12.9% to 26.0% of emergency physicians received one or more general payments each year. (Table 1) The number of emergency physicians accepting general payments was the highest (13,126; 26.0%) in 2015 and then constantly decreased to 9,935 (19.7%) in 2019. Similarly, the median annual total payments per physician decreased from $54 in 2014 to $47 in 2018, but increased in 2019. The number of emergency physicians accepting general payments significantly decreased each year between 2014 and 2019, with the relative annual percentage change (RAPC) of −2.9% (95% CI: −3.2 to −2.5, p<0.001), while there was no significant change in the per-physician total general payments. (Table 4)

When the COVID-19 pandemic occurred in the US in 2020, the total annual amounts, number of general payments, and the number of emergency physicians accepting payments substantially decreased between 2019 and 2020. The general payments per physician and the number of physicians accepting general payments decreased by 45.4% (95% CI: −57.7 to −29.6, p<0.001) and 47.8% (95% CI: −49.8 to −45.6, p<0.001) in 2020 compared to those between 2014 and 2019. (Table 4) During the COVID-19 pandemic period between 2020 and 2021, the per-physician general payments and the number of emergency physicians accepting general payments increased by RAPCs of 25.7% (95% CI: 5.9–49.2, p=0.009) and 21.7% (95% CI: 19.8–24.8, p<0.001), respectively.

Annual total research payments ranged from $29,602,868 in 2018 to $56,888,938 in 2021. (Table 3) Meanwhile, the number of emergency physicians receiving research payments was the lowest at 188 (0.4% of all emergency physicians) in 2021 and the highest at 304 (0.6%) in 2016. There were no clinically meaningful changes in research payments between 2014 and 2019. (Table 4) Also, there were no changes in research payments before and during the COVID-19 period, but the per-physician research payments significantly increased by 69.4% (95% CI: 28.9–122.7, p<0.001) in 2021 compared to those in 2020. (Table 4)

**Table 3.** Research payments to emergency physicians between 2013 and 2021.

| Variables | Program year |  |  |  |  |  |  |  |  | Overall |
| --- | --- | --- | --- | --- | --- | --- | --- | --- | --- | --- |
|  | 2013<sup>†</sup> | 2014 | 2015 | 2016 | 2017 | 2018 | 2019 | 2020 | 2021 |  |
| Total payments |  |  |  |  |  |  |  |  |  |  |
| Payment amounts, \$ | 11,063,957 | 46,283,114 | 50,728,394 | 53,123,761 | 45,116,819 | 29,602,868 | 32,481,344 | 37,374,351 | 56,888,938 | 362,663,547 |
| Number of payment contracts, n | 1,282 | 8,555 | 8,904 | 4,123 | 3,146 | 2,653 | 2,927 | 2,509 | 65,440 | 99,539 |
| Number of physicians with specific payment amount, n (%) | 114 (0.2) | 235 (0.5) | 279 (0.6) | 304 (0.6) | 283 (0.6) | 230 (0.5) | 218 (0.4) | 207 (0.4) | 188 (0.4) | 663 (1.3) |
| Payments per physician, \$ | | | | | | | | | | |
| Median (IQR) | 15,386<br>(4,834–101,900) | 19,541<br>(3,676–95,671) | 25,798<br>(7,000–95,870) | 25,752<br>(8,730–75,740) | 22,364<br>(5,080–88,625) | 25,782<br>(6,502–118,337) | 33,503<br>(8,081–122,299) | 41,725<br>(12,320–127,927) | 44,943<br>(11,453–200,961) | 62,843<br>(10,320–273,285) |
| Average (SD) | 97,052<br>(358,714) | 196,949<br>(667,820) | 181,822<br>(571,058) | 174,749<br>(790,249) | 159,423<br>(516,423) | 128,708<br>(357,725) | 148,997<br>(373,344) | 180,552<br>(449,742) | 302,601<br>(747,921) | 547,004<br>(1,884,010) |
| Range | 155–3,762,707 | 0.1–7,620,044 | 5–5,044,803 | 34–7,967,500 | 20–5,584,456 | 1–3,693,1101 | 12–4,111,636 | 105–3,448,920 | 12–5,391,808 | 0.1–22,538,680 |
| Gini index | 0.999 | 0.999 | 0.999 | 0.999 | 0.999 | 0.999 | 0.999 | 0.999 | 0.999 | 0.998 |

\*p<0.05, \*\*p<0.01, \*\*\*p<0.001
Abbreviations: interquartile range (IQR), SD (standard deviation)
<sup>†</sup> The payment data in 2013 were payments made to physicians between August 1 and December 30.

**Table 4.** Trends in general and research payments between 2014 and 2021.

|  | Relative annual average change rate (95% confidence interval), % |  |  |
| --- | --- | --- | --- |
|  | From 2014 to 2019 | 2014-2019 vs 2020-2021 | From 2020 to 2021 |
| General payments |  |  |  |
| Number of physicians receiving payments | -2.9 (-3.2 – -2.5)*** | -47.8 (-49.8 – -45.6)*** | 21.7 (19.8 – 24.8)*** |
| Payments per physician | 0.3 (-3.9 – 4.6) | -45.4 (-57.7 – -29.6)*** | 25.7 (5.9 – 49.2)** |
| Research payments |  |  |  |
| Number of physicians receiving payments | -2.8 (-5.3 – -0.1)* | -5.2 (-20.3 – 12.8) | -6.6 (-17.0 – 5.1) |
| Payments per physician | -8.8 (-17.8 – 1.2) | -27.6 (-56.4 – 21.4) | 69.4 (28.9 – 122.7)*** |

\*p<0.05, \*\*p<0.01, \*\*\*p<0.001
Payments for acquisitions, debt forgiveness, current or prospective ownership or investment interest, and royalty or license were excluded from the trend analysis of general payments.

## Discussion

To the best of my knowledge to date, this study is the first longitudinal analysis of both non-research and research financial relationships between emergency physicians and the healthcare industry using the Open Payments Database since the inception of the database in the US. The study brought several novel and important insights into the emergency physicians-industry financial relationships. This study illustrated that 56.8% of all active emergency physicians received 420,967 industry payments totaling more than $457 million from the healthcare industry in the US between 2013 and 2021. About one fifth of overall industry payments were made to emergency physicians as general payments, more than 70% of which were for speaking compensations for non-CME events and consulting fees. The very small number of emergency physicians received considerable amounts of general payments. While the payment amounts did not change, the number of emergency physicians accepting general payments significantly decreased since the inception of the Open Payments program in the United States. Additionally, only 1.3% of all emergency physician received research payments from the healthcare industry over the nine years period. There had been no change in research payments since the inception of the Open Payments Database, but the COVID-19 pandemic brought the substantial amounts of research payments to the very small number of emergency physicians.

There were differences in results between this analysis and previous studies. The findings showed that lower proportion of emergency physicians receiving payments but the per-physician payments was higher than those in previous studies.[5,6] Two previous studies reported that 30.1% to 35.4% of all emergency physicians received $18 to $44 in median total general payments per physician.[5,6] However, the study found that 12.9% to 26.0% of emergency physicians accepted $31−$54 in median annual general payments. This difference in the proportion of emergency physician with payments could be due to the difference in the data sampling methods between the previous studies and this study. The previous studies estimated the proportion of emergency physicians receiving payments from the Association of American Medical College Physician Specialty Data Report, while this study included all emergency physicians registered in the NPPES database. As Ahlawat et al. suggested, the NPPES database might have included larger number of physicians and might underestimate the proportion of physicians receiving payments. Also, as for the difference in payment amounts, the Open Payments Database includes many errors in specialty categorization in payment database. Even when payments were made to a same physician in one year, the several different specialties were assigned to the physician receiving payments in the payment database, as previously observed in rheumatology, allergology, and infectious diseases.[7,8,14] This difference in physician specialty could be caused by payment data being submitted by each company making a payment. Rigorous data collection process of this study could extract all payments made to the emergency physicians using the NPPES database and minimize the possibility of underestimation of per-physician payments.

Through the rigorous methodology, this study elucidated that the emergency physicians were the specialists with the lowest general payments ($31−$54 in median annual per-physician payments) made by the healthcare industry over the nine years. For other specialties, the median annual general payments per physician were $94 in pediatricians,[2] $140 in obstetrics and gynecology,[18] $197–$220 in infectious diseases,[14] $395−$597 in allergology and clinical immunology,[7] $407 in neurology,[19] $579−$616 in cardiology,[12] $648–$795 in oncology,[20] $500–$812 in rheumatology.[8] This finding was consistent with previous studies.[2,5,6] As Fleischman et al. pointed out, the healthcare industry has less incentive to make general payments to emergency physicians, as emergency physicians often provide short-term prescriptions and less expensive pharmacotherapy compared to other specialists.[5]

Notably, there were steadily decreasing trends in the number of emergency physician accepting general payments before the COVID-19 pandemic. In addition, the COVID-19 pandemic has significantly restricted the financial relationships between emergency physicians and the healthcare industry. Although the Open Payments Database was created to improve the transparency in the physician-industry financial relationships, it was also expected to decrease the influence of the healthcare industry on physician practices and patient care through public scrutiny and pressure on that relationship.[4,8,21] While the industry payments had been decreasing since the inception of the Open Payments Database in entire, but the increasing trends were recorded in several specialties such as oncology,[22] allergology,[7] rheumatology,[8] and infectious diseases.[14] Considering that even a small food and beverage payment significantly influence physician prescribing patterns[23-25] and substantial amounts of these payments and gifts sometimes made for promotions of drugs with less safety and lower efficacy with invalid evidence,[23,24,26] all physicians including emergency physicians should decline accepting the industry payments and gifts made for marketing purposes, even though physicians believe those financial interactions are beneficial and inconceivable to influence their clinical practice.[27]

Although the research payments included not only payments directly made to the emergency physicians but also costs for drugs, medical devices, and experimental equipment, inclusion of all research payments of which principal investigators were emergency physicians elucidated that the healthcare industry made nearly 80% of their expense to emergency physicians for research purposes. Additionally, only 1.3% of emergency physicians received those substantial research payments. Considering that, in previous studies conducted using the same methodology as this study, the percentage of physicians receiving research payments was approximately 7% in urology[28] and 11% in neurosurgery,[29] the research payments disproportionately concentrated on a much smaller number of physicians in the emergency medicine field. While there was increasing trend in research payments after 2020, these increased research payments were made to trials developing treatments for COVID-19 and the number of emergency physicians did not substantially change over the nine years. Therefore, the research payments from the healthcare industry could have temporarily increased and will decrease after the pandemic.

This study has a number of limitations. First, despite the physicians can review and dispute the payment data listed in the Open Payments Database, possible inaccuracy in the Open Payments Database could not be excluded as indicated by previous studies.[11,19] Second, the NPPES physician specialty was determined by self-declaration of each physician. As the methodological difference of physician specialty categorization between the NPPES database, the American Medical Association Physician Masterfile, and the Association of American Medical College Physician Specialty Data Report which is creased from American Medical Association Physician Masterfile database, the number of emergency physicians might be overestimated, as pointed out by Ahlawat et al.[19] Third, there would be unmeasured confounding factors influencing the amounts of and trends in industry payments such as physician age, position in their affiliation, career status, and race/ethnicity, and approval timing of novel drugs in emergency medicine of which information was not available through the NPPES database and the Open Payments Database.

## Conclusion

Majority of active emergency physicians received the general payments from the healthcare industry in the United States since the inception of the Open Payments Database. Only a small number of emergency physicians received considerable amounts of research and general payments. The general payments to the emergency physicians have been decreasing since the inception of the Open Payments Database in 2013, while the research payments were increasing due to the development of COVID-19 treatments.

## Data Availability

All data produced in the present study are available upon reasonable request to the author.

https://openpaymentsdata.cms.gov/

